# COVID-19 FATALITY RISK: WHY IS AUSTRALIA’S LOWER THAN SOUTH KOREA?

**DOI:** 10.1101/2020.05.14.20101378

**Authors:** Peter Collignon, John J. Beggs

## Abstract

**Background:** As the Covid-19 virus epidemic spreads, it is important to establish reliable estimates of fatality hazard rates. Australia and South Korea are ideal candidates for detailed consideration. Both have completed the first wave of the epidemic, they have extensive Covid-19 testing and tracking programs so that confirmed case load data are reliable, and neither country has had any significant case load stress in their hospital systems.

**Methods:** For each country, mortality hazard models were estimated using a parameterized distributed lag model where the number of daily deaths was dependent on the number of confirmed cases in each of the preceding six weeks. Age cohort CFRs were also examined.

**Findings:** We observed major difference in the mortality rates when comparing South Korea to Australia in both the simple age adjusted fatality rates and in the disease hazard curve. On a like-for-like basis, the CFR for South Korea appears to be close to double the Australian rate (aggregate; 2.4% vs 1.4%).

**Interpretation:** Neither differences in the time pattern of the peaking of the case load of confirmed cases, nor differences in the size of age cohorts of confirmed cases explain the difference in mortality observed. We discuss possible explanations that point the way for further investigation.

**Funding:** nil.

## INTRODUCTION

Reported Covid-19 fatality rates (CFRs) vary widely around the world (1). A major factor can be the different age profiles in different countries. In Singapore, the CFR is 0.1% but that is mainly because a large proportion of their cases are relatively young migrant workers (1,2), and that is in stark contrast to Italy with a CFR of 14.0% and UK with 14.3% (1). Despite the heterogeneous reported global data, it is remains important to establish dependable estimates of fatality hazard rates. Reliable data are necessary for calibrating the public health response, and for comparing clinical outcomes among institutions and among regional settings.

Several factors influence reported fatality rates. High levels of community spread can potentially impair the ability of the health systems to cope with the numbers of patients needing care, and hence a genuine rise in fatality risk. Locations that have limited testing capability are likely to see higher CFRs as only the most severe cases are identified and the denominator in the CRF ratio remains understated. Survival time from case confirmation to death is likely to be shorter when cases of infection are confirmed only in advanced stage of the disease, for example at the time of hospitalization. In the earliest stages of the epidemic, there may be a lack of awareness of the need to test for the disease, or there may not be suitable available tests to diagnose the disease. In this situation, both the number of infected cases and the number of deaths attributable to the disease may be understated.

The first objective of this study was to estimate the daily death hazard rate from time of case confirmation to case resolution. In order to get a more reliable estimate the paper uses data from two countries, Australia and South Korea. Both these countries have extensive Covid-19 testing and tracking programs, and both have not had any significant case load stress in their hospital systems from Covid-19. Their crude mortality rates are 1.4% and 2.4% respectively. Both countries have been able to reduce their ongoing new cases to relatively small numbers. This means the data form these two countries are close to a complete picture of a first wave of the epidemic.

The estimated hazard function can also be used to estimate the terminal case fatality rate, CFR^Terminal^. This estimate corresponds to the retrospective CFR if the disease were eradicated. The purpose of the calculation is to remove a bias in the reported CFR that occurs because at any point in time the case load contains a backlog of, as yet uncounted, individuals who will in due course due die from the disease.

## METHODS

### Sources of data

Data on daily number of confirmed cases and deaths were obtained from the Australian Government Department of Health web sites (3) and from the Johns Hopkins Coronavirus Resource Center for the period January 22^nd^ to May 12^th^ 2020 (1).

### Statistical analysis

For each country we used a parameterized distributed lag model where the number of daily deaths was dependent on the number of confirmed cases in each of the preceding six weeks. Denote the hazard function as,

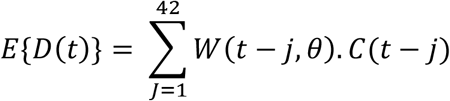

where *E*{*D*(*t*)} is the expected number of deaths on day t, *C*(*t*−*j*) is the number of confirmed cases j days earlier, *W*(.) is a vector function that acts as a set of weights what proportion of cases of confirmed cases can expect to die j days after case confirmation. The vector *θ* holds the parameters of the weighting function. *W*(.) will be modelled as a simple linear function,

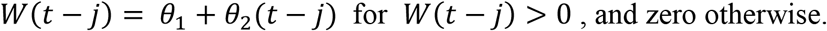

The linear weighting function must only be regarded as an approximation of reality, but it does have the advantage that it is parsimonious in the number of parameters that need to be estimated, and the shape of the function is transparently obvious. When longer and more detailed data sets eventually become available, more complex weighting schemes such as Weibull distributions may provide more generalized insights.

The number of deaths on any day, *D*(*t*), was modelled as a draw from a Poisson distribution with expected number of deaths *E*{*D*(*t*)}. The model’s parameters were estimated by maximum likelihood and results reported below.

Materially similar estimation results to those of maximum likelihood can be obtained by applying non-linear least squares estimation over the sum of the squared difference between actual deaths, *D*(*t*), and modelled deaths, 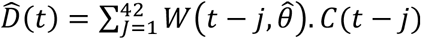 or alternatively over the sum of the squared difference between the cumulative sums of actual and modelled deaths.

## RESULTS

Age specific CFRs for Australia and South Korea show the same general pattern of higher mortality among the older cohorts, and minimal mortality risk at younger cohorts. Table 1 shows age specific CFRs are higher for Korea and Australia in all age cohorts, and in aggregate. This knowledge leads us to believe that the daily death hazard rates for South Korea will be above those for Australia.

**Table 1.** Case Fatality Rates by Age Band.

| Age Band | South Korea | Australia |
| --- | --- | --- |
| 80+ years | 25.3% | 20.1% |
| 70-79 years | 10.8% | 4.0% |
| 60-69 years | 2.7% | 0.7% |
| 50-59 years | 0.8% | 0.2% |
| 40-49 years | 0.2% | 0.1% |
| 30-39 years | 0.2% | 0.0% |
| 20-29 years | 0.0% | 0.0% |
| 10-19 years | 0.0% | 0.0% |
| 0-9 years | 0.0% | 0.0% |
| Aggregate | 2.4% | 1.4% |

Timing differences in case onset can play a role in explaining reported CFRs. Countries that have earlier experience of the virus, such as South Korea, will have a larger proportion of resolved case. Countries that have later experience of the virus will have relatively more unresolved cases, some of which will subsequently die, causing their reported CFRs to be biased downwards.

Numerical estimation by maximization of the Poisson likelihood function converged rapidly, and the coefficients of the fitted hazard function are shown in the Table 2 below. The estimated parameters are highly statistically significant, implying that the model provides a strong fit to the data. Figures 1 and 2 below provide a visual sense of how well the models track the data.

**Table 2.**
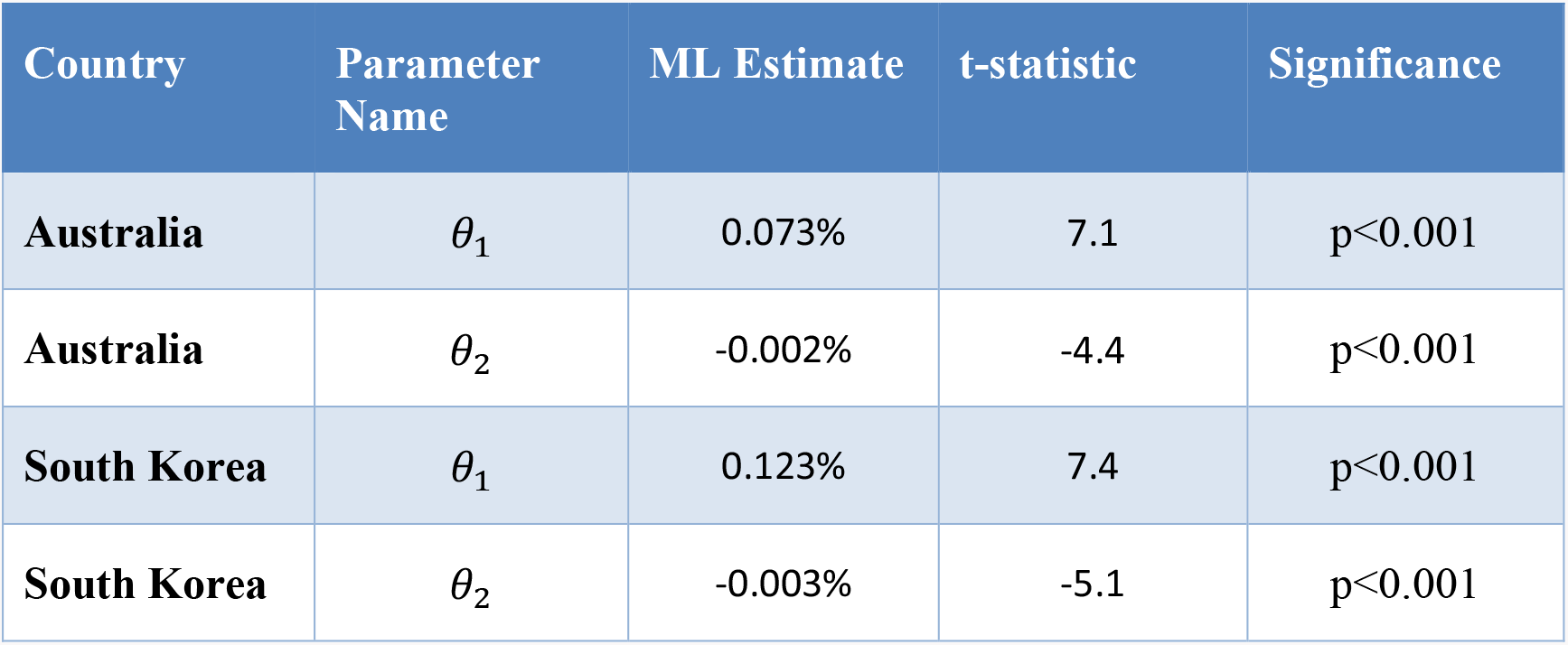
Model Parameter Estimates.

**Figure 1.**
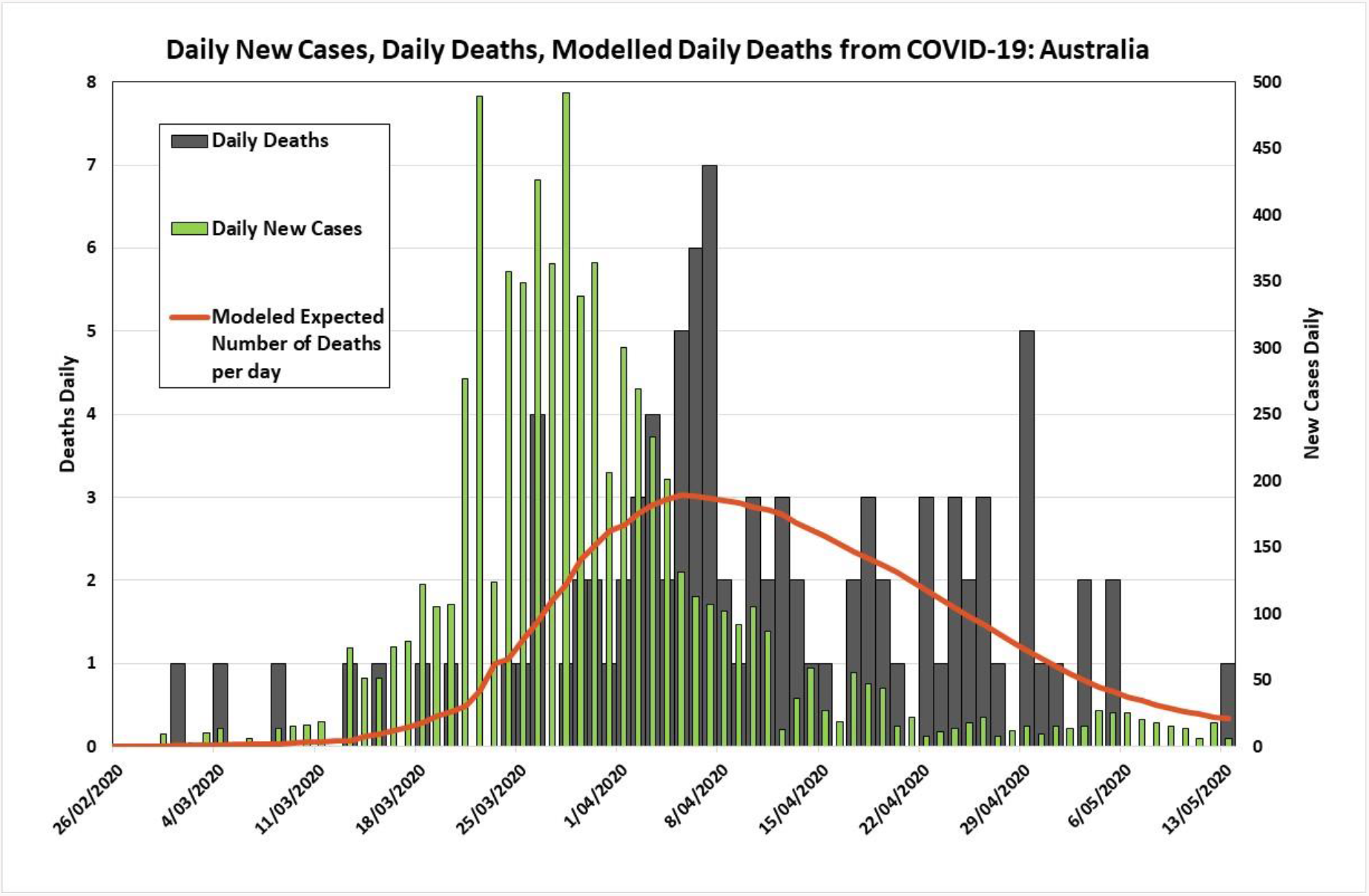

**Figure 2.**
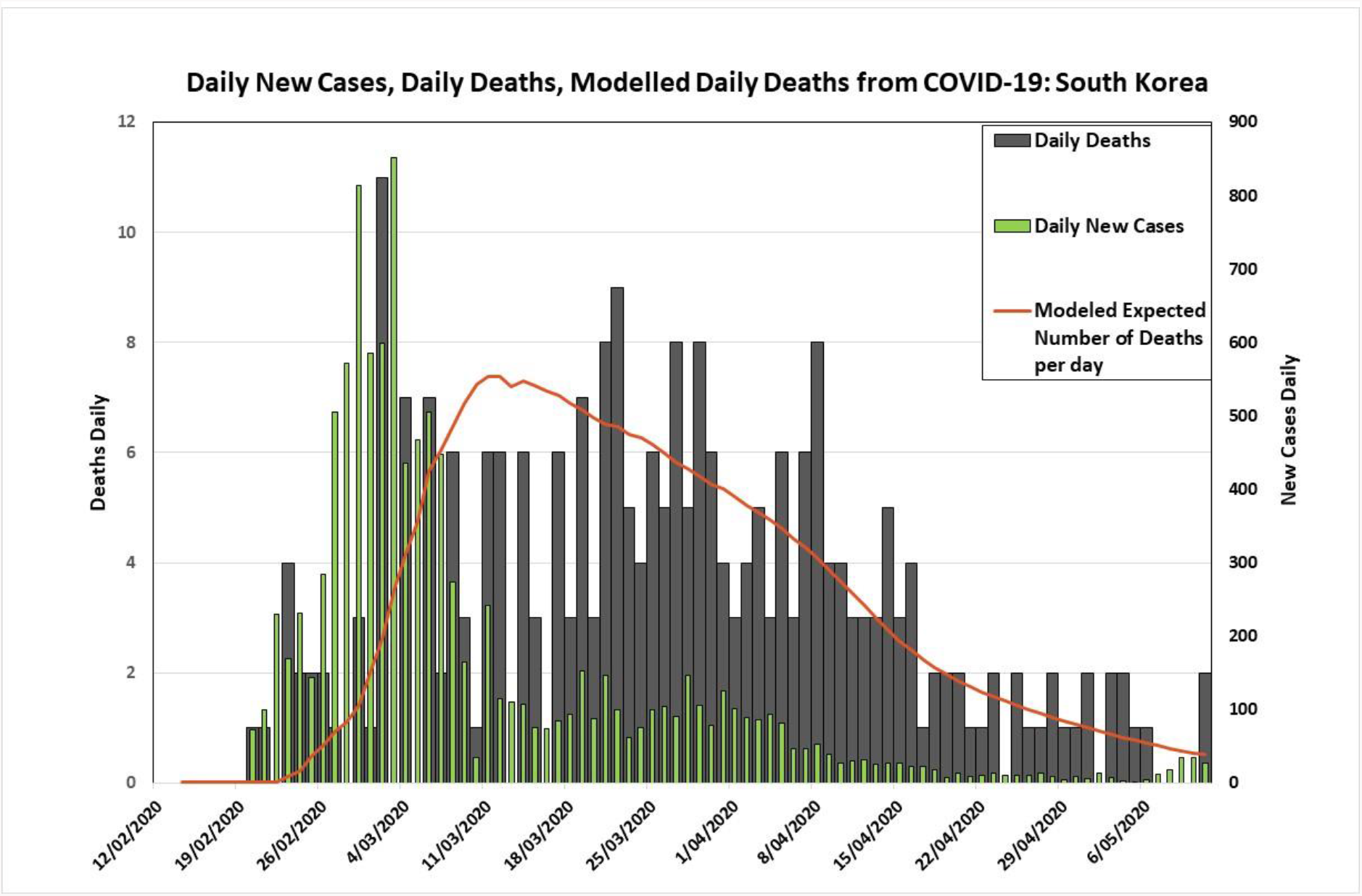

Health authorities in both South Korea and Australia were able to test early and extensively for Covid-19, but it needs to be noted that relatively much less testing per capita was carried out before the first few virus fatalities. This fact is apparent in Figures 1 and 2 where the fitted models under-estimate the number of deaths at the very start of the epidemic.

Visually, the estimated models shown in Figures 1 and 2 display good agreement with the fatality data. The case data shows a lot of day-to-day variability, but the peak in deaths appears to lag the peak in confirmed cases by a range of 10 to 14 days.

Figure 3 show plots of the estimated hazard function for both countries, and it also provides the asymptotic 95% confidence intervals derived from the maximum likelihood estimation.

**Figure 3.**
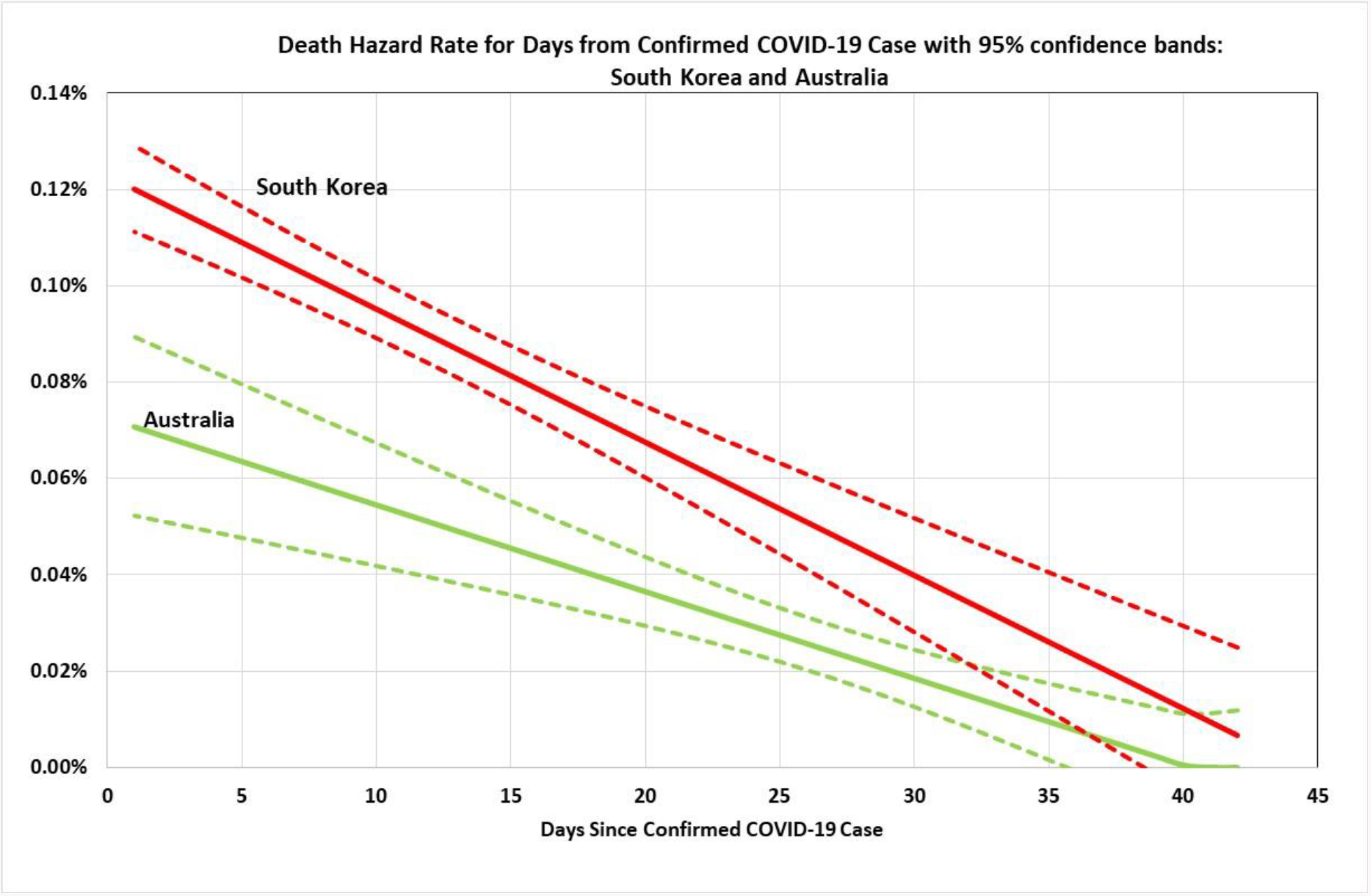

Figures 4 and 5 plot the historical reported CFRs for each country. The solid line show a pattern of initially falling CFRs due to the initial under identification of cases at the start of the epidemic, discussed above in the context of Figures1 and 2. The broken lines show the projected evolution of the CFRs if each country ceased having any more covid–19 cases at the 7^th^ May, and the current case load was held through to resolution. The dotted lines asymptote are the terminal values of CFR^Terminal^ of 1.46% for Australia and 2.43% for South Korea. Differences in CFR^Terminal^ reflect fundamental difference the counties’ fatality rates after removing the case resolution effects due to timing differences in the arrival and evolution of the epidemic.

**Figure 4.**
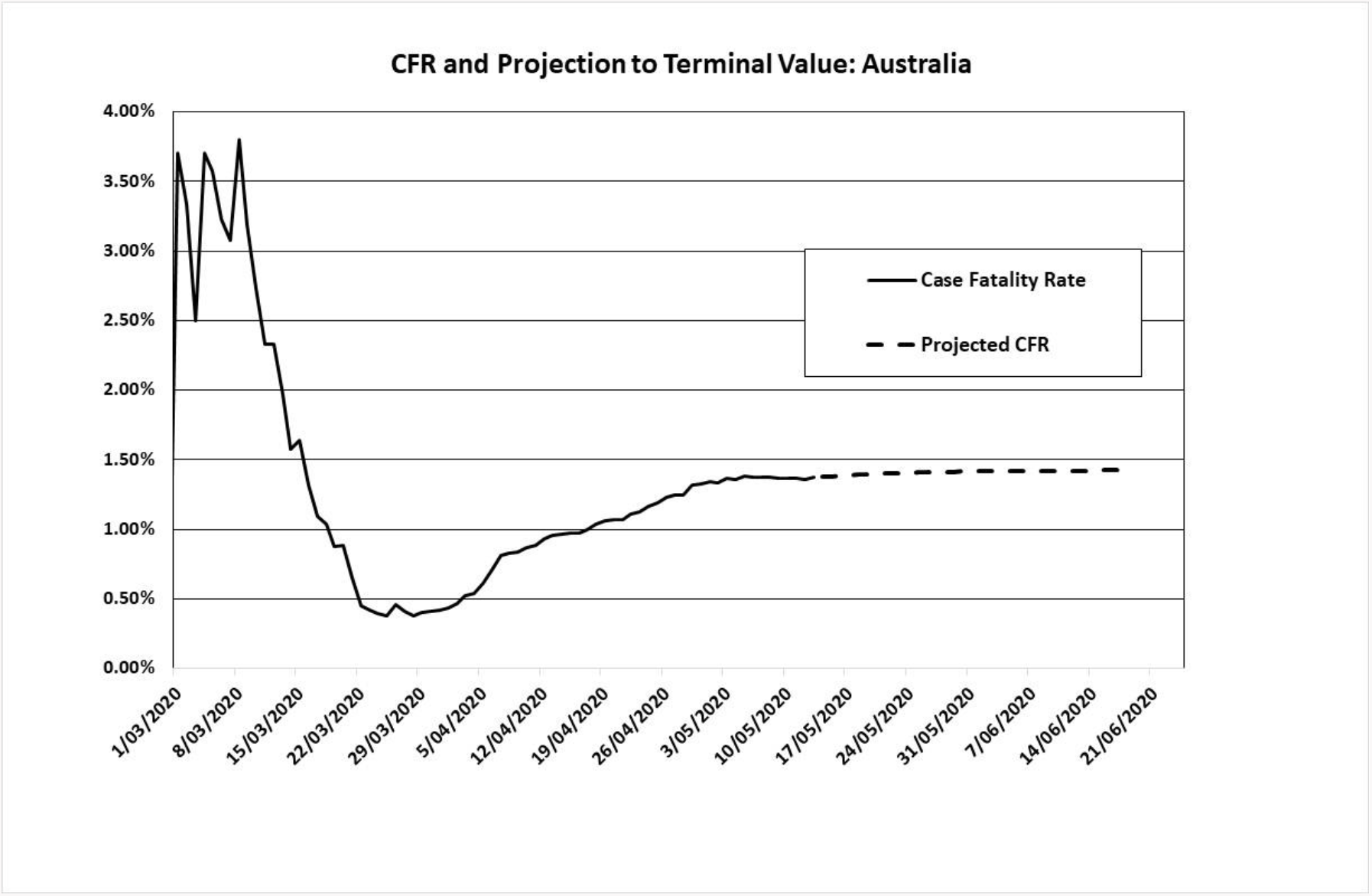

**Figure 5.**
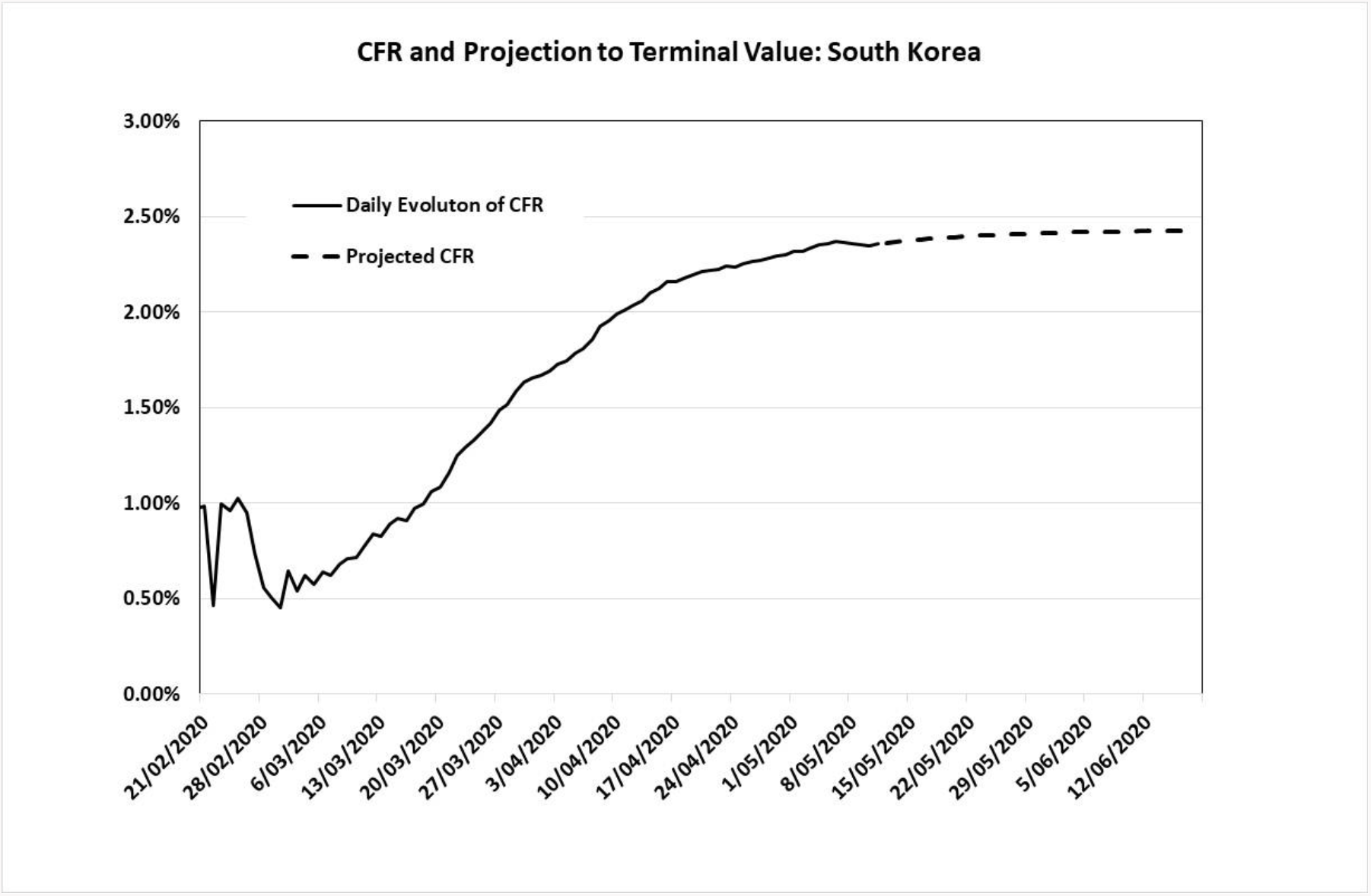

## DISCUSSION

When the age specific CFRs from South Korea are applied to the age profile of the Australian case load experience as at the 9th May, they predict 265 deaths, and an aggregate Australian CFR of 2.6%. This CFR is slightly higher than the actual Korean CFR of 2.4% which reflects the fact that Australia virus victims are more concentrated in older age groups than in Korea. But this projected fatality rate of 2.6% is nearly double the actual Australian CFR of 1.4% at May 9^th^.

Figure 3 displays that the hazard rate curve for Australia is significantly below that for South Korea (formally, p<0.001), with terminal case fatality rates, CFRTerminal, of 2.43% and 1.46% respectively. These results confirm that differences in the aggregate reported CFRs for these two countries cannot be explained by the time pattern of the peaking of confirmed cases in each country. Since both countries have advanced high-quality health systems, and since the epidemic in both countries was quickly contained, and each country’s hospital infrastructure was not overwhelmed, it remains unclear what accounts for this difference in fatality rates.

An interesting observation (see Figures 4 and 5) is that the case fatality rates in Australia were much higher in the initial couple of weeks of the epidemic (initially over 3% and later 1.4%). This phenomenon is well known and observed in other countries, and it is most likely related to the lower counting number of confirmed cases identified early in the epidemic (4). Another important factor in Australia was the disproportionately larger number of cases involving older age groups returning from travel abroad, particularly from cruise ships.

Korea was in winter while Australia was in late summer and autumn when the epidemic arrived. It might be that other viruses and bacteria (e.g. pneumococcus) spread more easily in winter and cause more complications in those that developed lung involvement. Variations in death rates were seen during the Spanish flu epidemics in 1918, and much of this variation was due to variations in rates of secondary infections and/or immunity to bacteria and viruses (5,6,7).

Minor differences in the strains of the virus that have infected each country may lead to variations in associated mortality. A further factor is that because Covid-19 is most dangerous for older people with co-morbidities, Koreans aged in their 70’s and 80’s, who were children at the time of the Korean War and just afterwards, may carry additional co-morbidities specifically due to nutritional and other legacy effects of that war-time experience [8]. Finally, there may be differences in standard hospital treatment regimens (including in ICU’s) that are important and which need to be identified. Korea also has higher rates of antibiotic resistance than Australia for many pathogens (9,10). If bacterial secondary infections contribute to deaths of people with Covid-19, this might be additional factor.

Australia had many sources for its cases of Covid-19, but a higher proportion than in Korea were related to overseas travel (mainly USA and Europe but with an equal proportion returning from cruise ships) and there was relatively little community transmission in Australia without a confirmed contact (10%) (3). This experience is similar to what occurred in Korea, with Europe and the Americas their main source for imported cases. But Korea’s large initial outbreak and then local transmission clusters were more likely related to strains from China (11). So, viruses more recently from people infected in China, likely contributed a larger proportion of their cases compared to Australia. This might be of significance if strains have changed in their pathogenicity in different regions with time.

The estimated hazard functions imply that deaths lag case confirmation by an average of 13.7 days in Australia and 15.1 days in Korea days. Another important observation is that the fatality pattern for Covid-19 appears to have quite a long tail. Although the disease can cause fatalities shorty after case confirmation, there remains a risk of a fatality occurring even more than a month after case confirmation. A factor in seeing a more prolonged time between case confirmation and death could be that when a health system is able to cope, patients will likely spend much longer times in ICU and with more interventions than when a health system is overwhelmed. If this is true, we may see shorter trailing fatalities when data becomes available for stressed hospital systems such as in Italy, the UK and New York State.

In conclusion, we have seen a major difference in the mortality rates when comparing Korea to Australia both in the disease hazard curve and also in the simple age adjusted fatality rates. It will be important to better explore difference in the virus strains, variations in carriage of bacteria (e.g. pneumococcus) and/or how healthcare is delivered to try and unravel what are the most important factors that may have contributed to this difference.

## Data Availability

All data for this paper is in the paper or can be found on the web by following the links in references

